# Patterns and Purpose of Social Media Use, and Their Association with Depression Among Students at Three Universities in Tanzania: A Cross-Sectional Study

**DOI:** 10.64898/2026.07.26.26357970

**Authors:** Nyakato Caroline Nshala, David Thomas Tarimo, Vanessa Andronis David, Secilia Alphonce Ntanga, Kim Madundo

## Abstract

Social media has an estimated global user base of 3.5 billion. Adolescents, constituting over 90% of this group, and thereby university students. are extensively involved in social media use, emphasizing its significance in their daily lives. Excessive use of social media poses risks such as depression, affecting academic performance.

The objective of this study was to determine the association between social media use and depression among university students in Urban Northern Tanzania.

This was a cross-sectional study that involved university students from three universities. Data was collected through an online close-ended questionnaire. Statistical analysis included the use of frequencies, percentages, chi-square and bivariate logistic regression at 95% confidence intervals (CIs) and significance at p-value <0.05.

A total of 384 participants were enrolled in the study. 36.7% reported that they used social media for 1 to 3 hours daily. 58.1% of all participants reported high frequency of social media use for social purposes. 29.2% of university students who participated in the study were screened to have depression. The prevalence of depression increased with longer daily duration of social media use. Those using social media for more than 5 hours daily, had 3.049 times higher odds of being depressed. Moderate frequency of social media use for social purposes was linked to reduced depression.

Daily duration of social media was significantly associated with depression levels among university students. Significant associations were also found between moderate use of social media for socializing, with lower risk of depression. We recommend that universities develop targeted mental health interventions and promote balanced, purposeful social media use among students.

## Introduction

Social media use is rapidly growing, with estimates of nearly four billion active users across the world (1). Up to 100% of university students regularly use social media depending on the setting (2,3). Social media platforms have become a crucial part of, and are seamlessly integrated into, students’ lives, offering avenues for connection, sharing information, entertainment, and education (4,5).

Social media usage patterns vary by country and setting. Globally, adolescents spend on average 2.3 hours per day using social media (6). Findings from sub-Saharan Africa indicate longer duration of daily use, up to 3.7 hours (7). In Tanzania, the daily duration of use is lower than this regional average, at approximately two hours (8). Studies on social media use among university students identified entertainment, communication, and seeking information as common reasons for use (2,4,9). In Saudi Arabia, less than 1% of university students utilize social media for academic purposes (10). In Bangladesh, the most common purpose (45%) of social media among university students was communication, with approximately a minority (10%) utilizing it for seeking and sharing information and knowledge (9). In contrast, the majority of university students in Nigeria used social media for seeking information; both news and for academic reasons (2). Similarly, the majority of university students in Kenya referred to social media for information and knowledge (56%), and half (51%) of the students socializing on these networks (11). In Tanzania, limited information is available on social media use among university students; however, communication was identified as the most common purpose of use (25%) among the general population (8).

In addition to the frequent, varied, and highly accepted use of social media by university students (2,4,9), concerns have emerged about its potential association with mental health challenges, particularly depression (1,11). Depression is one of the most common mental health challenges worldwide (12), and its onset is three-times as common during adolescence – which coincides with university age – compared to other age groups (13). The prevalence of depression among university students varies widely by country and setting, with findings ranging from 4% to 57% (14–16).

The mechanisms of association between social media and depression are varied. Studies have identified higher durations of social media use, purpose of use, and the type of platform used as predictors of depression (1,17).

Furthermore, as access to social media increases in Tanzania (8), depression remains under-detected (18,19). Previous literature highlights that nearly half of individuals with depression do not have an established diagnosis (20). Further, depression in university students is associated with negative outcomes including decline in academic performance, poor academic engagement, dissatisfaction with academic life, dropout, and suicide (21–23).

Despite increasing awareness of the relationship between social media use and depression among university students, there is a lack of literature on how specific patterns of social media use may be associated with depression, particularly within a resource-limited context such as Tanzania. This research aims to address this knowledge gap by exploring patterns of social media use, depression and their association among university students at three universities in northern Tanzania.

## Materials and methods

### Study design

The reporting of this study has followed the STROBE guideline (24).

An analytical cross-sectional study was conducted among university students from three universities in Moshi Municipal in Northern Tanzania; all data was collected from April 1^st^ to June 30^th^, 2024.

### Study setting

The three study sites were; Kilimanjaro Christian Medical Centre University (KCMCU), Moshi Co-operative University (MoCU), and Mwenge Catholic University (MWECAU). These institutions were selected to represent diverse academic disciplines, campus environments, and student demographics. The universities are all located in an urban setting, and all within a 10-kilometre radius. KCMCU is a health and allied sciences private institution, offering courses on Medicine, Pharmacy, Medical Laboratory Sciences and Nursing. At MoCU, a public university, courses include various social, business, and technology-related courses: Accounting and Finance; Business Administration; Community and Economic Development; Economics; Human Resource Management; and Information, Communication and Technology. At MWECAU, a faith-based private university, courses include science, business, social and Law related courses: Chemistry, Computer science, Applied Biology, Mathematics and statistics, Education, Accounting and Finance, Procurement and Supply chain Management, Project Planning and Management, Business Administration Management, Social Work and Human rights, and Law.

### Study population

Eligible participants were all diploma- and undergraduate-level students enrolled at either KCMCU, MoCU, or MWECAU at the time of the current study, aged 18 and above, and able to provide informed consent. Students who did not own smartphones or personal computers, who were on personal leave of absence, or who were unable to participate due to physical or mental reasons were excluded.

### Sample size and sampling technique

We referred to Kish Leslie’s formula for cross-sectional studies, N= [p(1-p) z^2^]/d^2^ (25), with an assumed proportion of 50% of the sample having depression. The minimum estimated sample size was 384 participants. We utilized a stratified sampling strategy, where each institution represented a stratum (26). Due to similarities in intake numbers at each of the universities, we intended to recruit one-third (approximately 33%) of the participants from each site. Thereafter, we used convenience sampling at each institution until the sample size was reached (27).

### Data collection methods, tools and procedures

We collected data using an online, self-administered, closed-ended Google form survey in English. Data was collected from 1^st^ April to 30^th^ June, 2024. The researchers visited each study site and met with class representatives to inform them about the study aims. The surveys were then shared with the overall student bodies via the class representatives. Study participants were able to access the study questionnaire through their smartphones or personal computers. The survey took approximately 10 minutes to complete and included four sections: demographic information; duration and forms of social media use; purpose of social media use; and a screener for depression. The survey explicitly made participants aware that this study was investigating the use of specific social media platforms and not merely the internet.

*Demographic data*; Independent variables included age group, gender, university, current level of enrollment (diploma or Bachelor’s level), and year of study.

*Duration, forms and purpose of social media use*; We incorporated a modified social networking usage scale by Gupta and Bashir (28). The tool comprises 20 items, grouped into four domains: *academic*, *socialization*, *entertainment*, and *informativeness*. Each item is scored on a 5-point Likert scale ranging from 1 (strongly disagree) to 5 (strongly agree), allowing for a total score between 20 and 100. Higher scores indicate greater perceived utility and engagement with social networking platforms. The scale was originally developed and validated in India among university students. Exploratory factor analysis confirmed its four-factor structure, and it demonstrated good internal consistency with a Cronbach’s alpha of 0.83. Although the Modified Social Network Usage Scale by Gupta and Bashir (2018) has not yet been formally validated in Tanzania, its applicability is supported by its use in a comparable East African setting (11). Given these parallels, the scale is expected to perform adequately in the current sample, pending future validation efforts. The tool was translated to Swahili by the first two authors and back-translated to English by the third and fourth authors. Revisions were made the study supervisor (final author). The tool was then pre-tested on a non-study sample of 10 people for comprehension.

*Depression*; The Patient Health Questionnaire 9-item version (PHQ-9) was used as a screener for depression (29). The tool has previously been translated to Kiswahili and validated in the Tanzanian context (30), and used in university student samples (31). In Tanzania, it has demonstrated very good internal consistency (Cronbach’s alpha = 0.83) (30). The PHQ-9 comprises nine items which match the Diagnostic and Statistical Manual of Mental Disorders version 5 (DSM 5) criteria for major depressive disorder (32). These symptoms are required to have been present for a minimum of two weeks’ duration, whereby each item can be scored from 0 (no symptoms at all) to 3 (symptoms nearly every day) with a maximum score of 27. The cut-off scores on this current study were derived from the earlier validation study in Tanzania, categorizing depression severity as follows: 0–4 (minimal), 5–9 (mild), 10–14 (moderate), 15–19 (moderately severe) and 20+ (severe symptoms) with a score ≥9 being equivalent to a major depressive episode, or clinically significant depression (29). Scores of 10 and above were classified as positive for depression in this study.

### Data management and analysis

Data was entered and cleaned by the first four authors. We used the Statistical Package for Social Sciences (SPSS) version 20 to analyze the data. Categorical variables were summarized using frequencies and percentages, while numerical variables were summarized using means and standard deviations. The Chi-Square test for independent association was used to assess whether there was a significant link between categorical variables and depression among the students. The strength of association between social media usage patterns and levels of depression was measured using odds ratio (OR) at a 95% Confidence interval (CI). A p-value of less than 0.05 was considered statistically significant. The final author supervised data entry, analysis and management.

### Ethical considerations

This research was guided by a strong commitment to ethical principles, ensuring the well-being, confidentiality, informed consent, anonymity and rights of participants. A Research Ethical Clearance Certificate with the number UG 54/2024 was obtained for ethical approval from KCMCU Institutional Review Board (IRB) called College Research Ethics and Review Committee (CRERC).

Participants were asked for written informed consent at the beginning of the questionnaire, with clear communication about the study’s purpose, procedures, and potential risks.

## Results

### Participants background characteristics

A total of 384 participants were enrolled in the study, with a slight majority being female (53.4%). The median age was 22 (SD ±3.267) years, predominantly within the 18-24 range (89.8%), typical for a young adult university population. Participants were distributed almost equally among the three universities: KCMCU (33.9%), MoCU (32.6%), and MWECAU (33.6%). Most were undergraduates (79.2%), with a smaller percentage in diploma programs (20.8%). The majority were in their second (33.1%) and third (34.4%) years of study, with fewer participants in the first, fourth, and fifth years. **(Table 1)**.

**Table 1:** Socio-demographic characteristics of study participants (N=384)

| Variable | Frequency | Percentage |
| --- | --- | --- |
| **Gender** |  |  |
| Female | 205 | 53.4 |
| Male | 179 | 46.6 |
| **Age** |  |  |
| Median (SD) | 22 (3.267) |  |
| 18 - 24 | 345 | 89.8 |
| 25 and above | 39 | 10.2 |
| **University** |  |  |
| KCMCU | 130 | 33.9 |
| MOCU | 125 | 32.6 |
| MWECAU | 129 | 33.6 |
| **Level of study** |  |  |
| Diploma | 80 | 20.8 |
| Undergraduate | 304 | 79.2 |
| **Year of study** |  |  |
| First year | 108 | 28.1 |
| Second year | 127 | 33.1 |
| Third year | 132 | 34.4 |
| Forth year | 8 | 2.1 |
| Fifth year | 9 | 2.3 |

### Social Media Platforms usage

Six social media platforms were listed. of which the majority, 91.7% (n= 352), reported using WhatsApp, 75% (n= 288) reported using Instagram, 45.1% (n= 173) reported using TikTok, 44.3% (n= 170) reported using Twitter (X), 26% (n= 100) reported using Snapchat, 22.7% (n= 87) reported using Facebook and 5.2% (n= 20) reported using other platforms besides the listed **(Fig 1)**.

**Fig 1.**
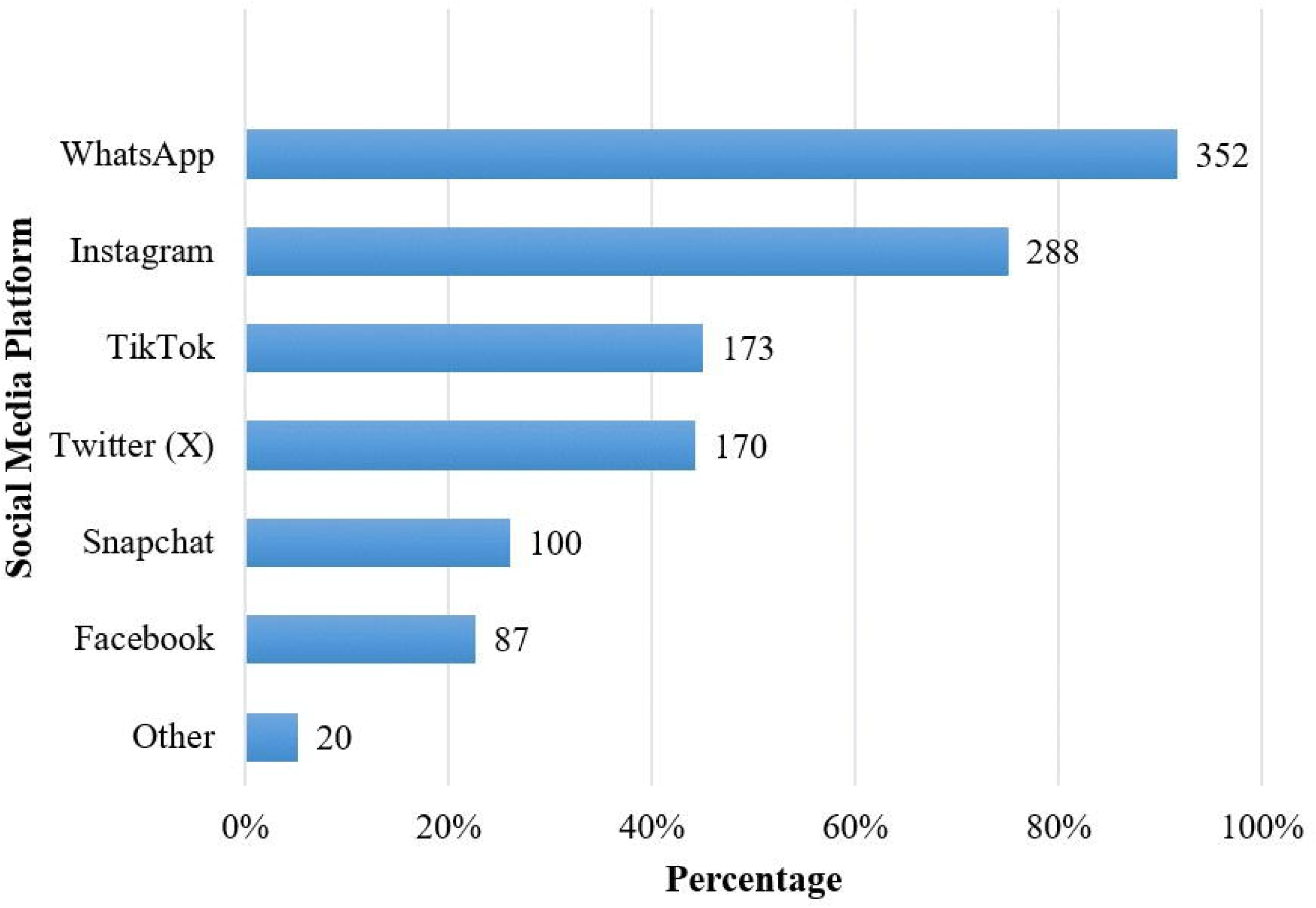
Social Media Platforms Usage (N=384).

### Daily duration of social media use

Among the 384 study participants, 36.7% (n=141) reported that they used social media for 1-3 hours daily, 29.7% (n=114) reported that they used social media for 3-5 hours daily, 18.8% (n=72) reported that they used social media for less than one hour daily and 14.8% (n=57) reported that they used social media for more than five hours daily **(Fig 2).**

**Fig 2.**
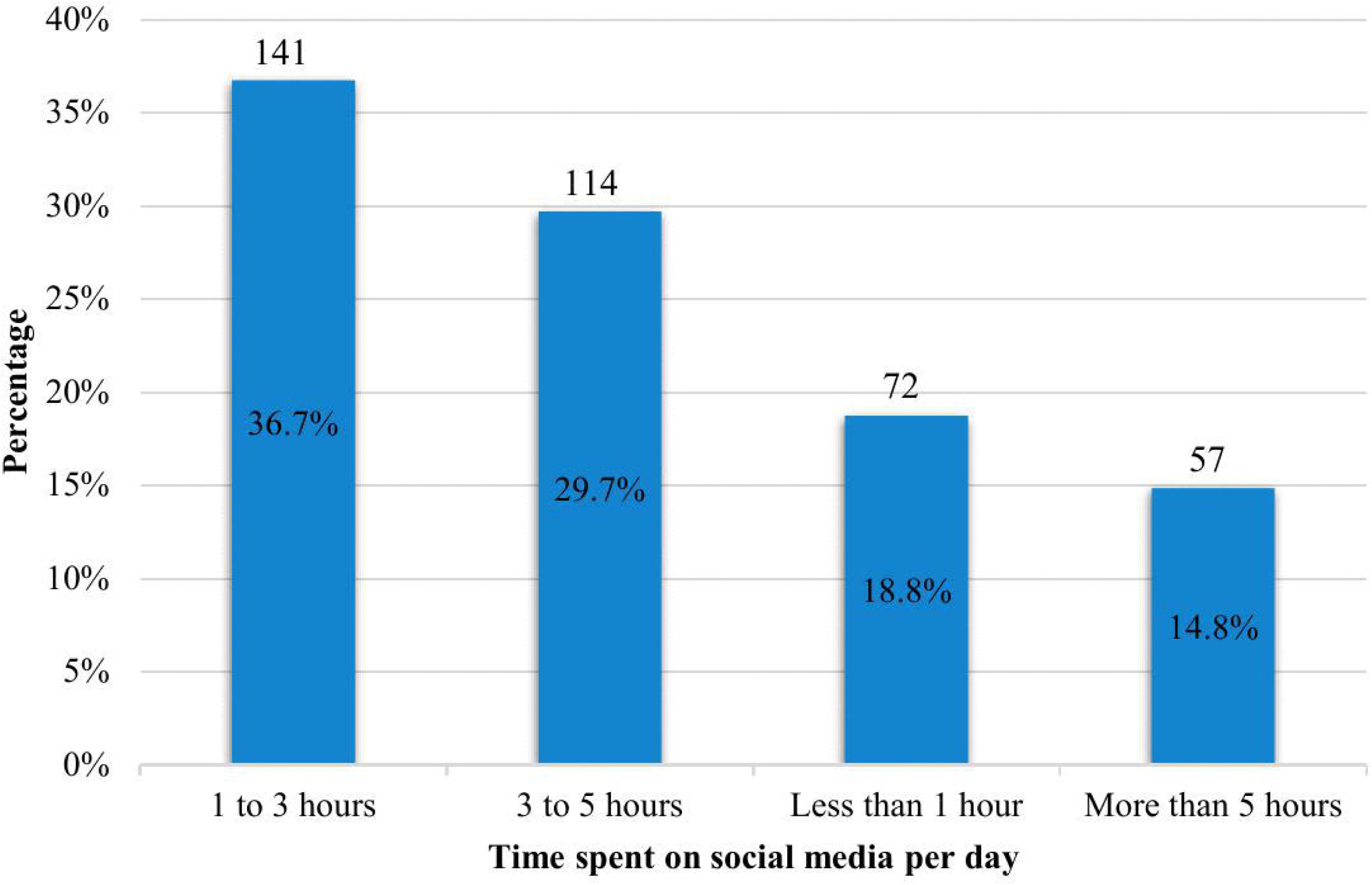
Daily duration of social media use (N=384).

### Purpose of social media use

Out of the 384 study participants, more than half (50.8%) reported a high frequency of use for social purposes. The majority (58.1%) also reported high frequency of use for academic purposes. Using social media as a source of entertainment was also common, with 172 students (44.8%) reporting high frequency of use and another 172 students (44.8%) reporting moderate frequency of use. Additionally, almost half of the students (47.7%) reported a high frequency of using social media to gain information **(Table 2)**.

**Table 2:**
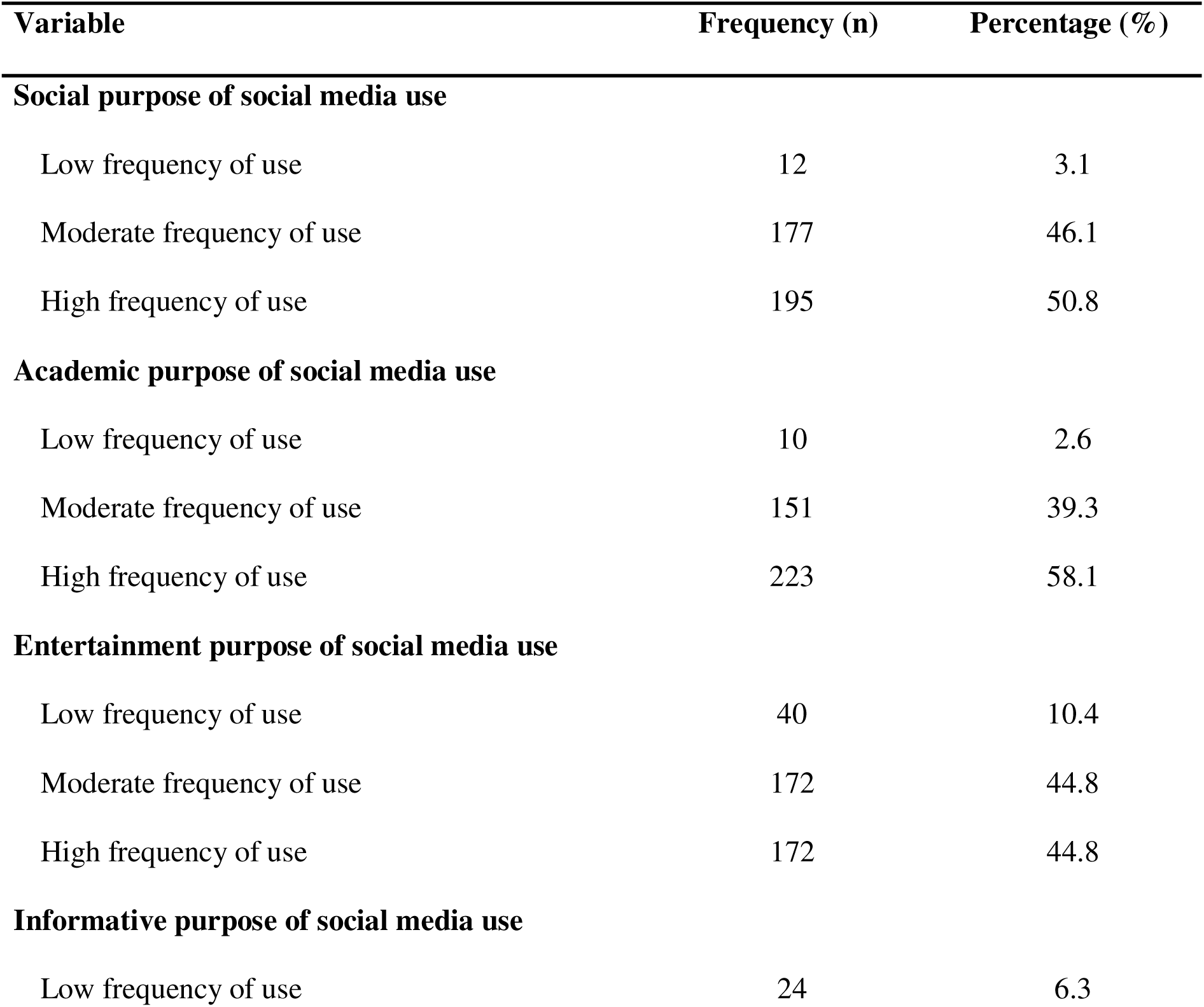
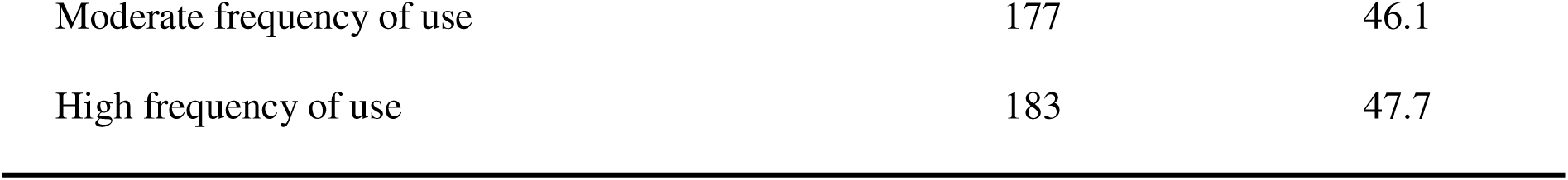
Purpose of social media use. (N=384)

### Prevalence of depression

Overall, a total of 112 (29.2%) individuals screened positive for depression, while 272 students (70.8%) self-reported no depression **(Fig 3)**.

**Fig 3.**
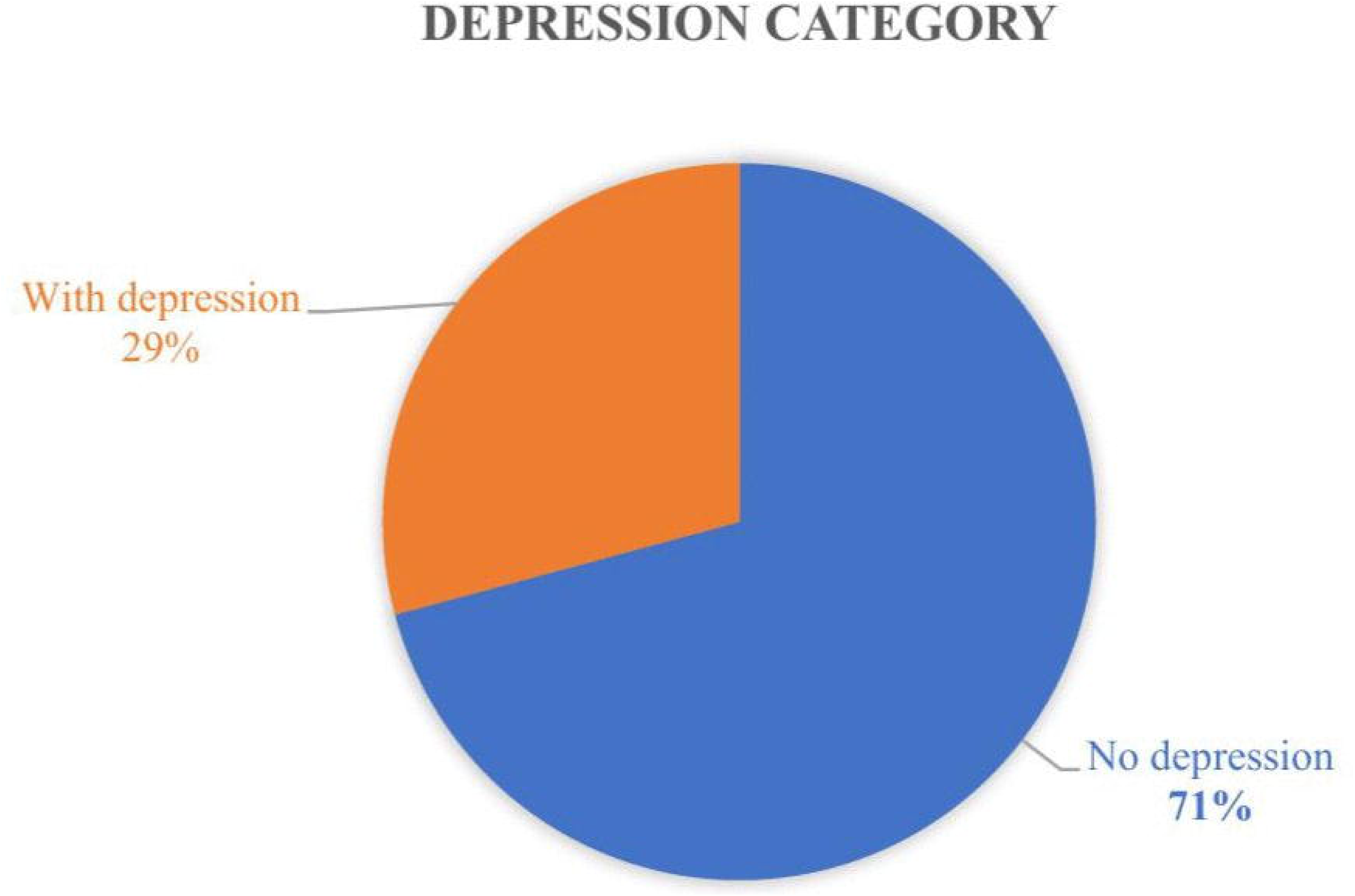
Category of level of depression (N=384)

Among 384 respondents, more than one-third of the students 38.5% (148) had a mild level of depression, 23.2% (89) had a moderate level of depression, 3.9% (15) had moderately severe depression, while 1.8% (7) screened positive for severe depression and the remaining one-third (32.6%) had no depression. The mean PHQ-9 score for depression was 7.16 (SD ± 4.771), demonstrating that on average, the students had a mild level of depression **(Table 3)**.

**Table 3:**
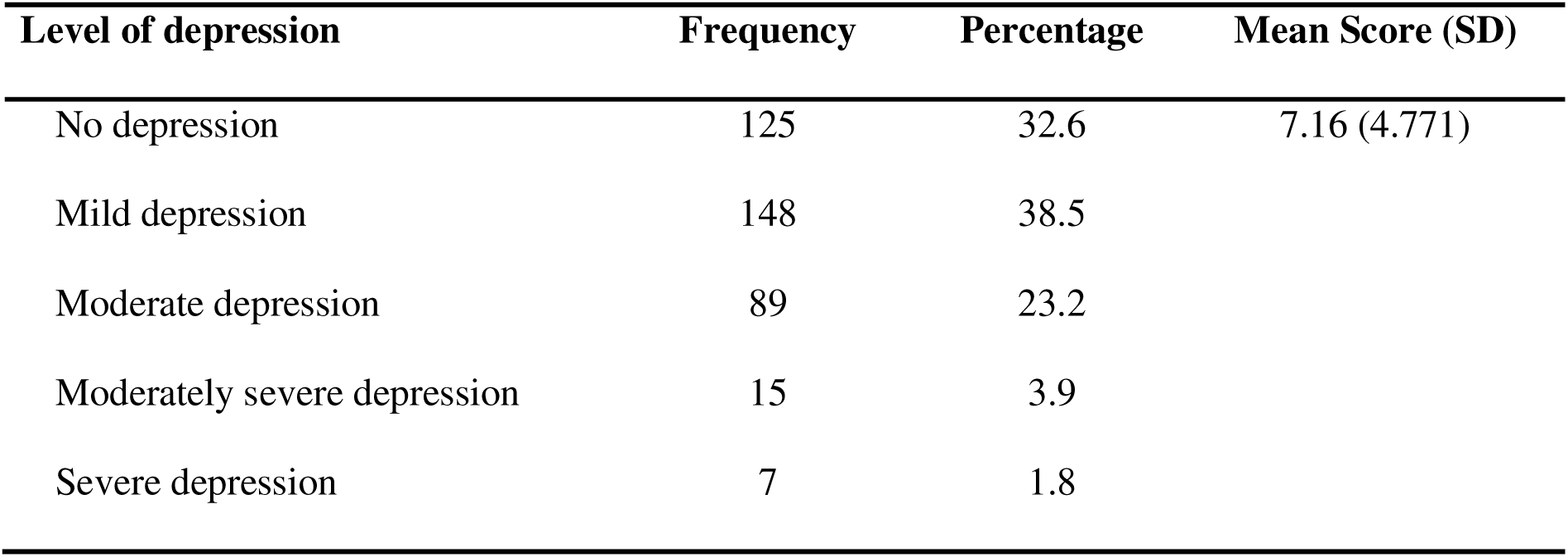
Levels of depression (N=384).

### Association between social media usage patterns and depression

Among 384 university students who participated on the study, the prevalence of depression increased with longer daily duration of social media use; less than one-quarter of participants (22.2%) who used social media for less than one hour per day reported depression. On the other hand, almost one-half of the students (49.1%) who used social media for more than 5 hours a day reported depression. The Adjusted Odds Ratios (AORs) indicated that compared to the reference group (less than 1 hour per day), the odds of depression were 0.773 times for those using social media for 1-3 hours daily (p= 0.497), 1.7 times higher for the 3-5 hours daily group (p= 0.146), and 3.049 times higher odds of depression among the >5 hours per day sub-group (p= 0.007).

Our findings regarding purpose of social media use revealed that students with moderate use of social media for social purposes were 78.9% less likely to have depression compared to students who reported low frequency of use for social purposes. (p= 0.026). Similarly, high frequency users of social media for social purposes were 51.4% less likely to be depressed compared to low frequency users (p = 0.310). Use of social media for academic purposes, both moderate and high frequency use, were associated with lower odds of being depressed compared to low frequency users, though not statistically significant (p= 0.53, p= 0.408, respectively). Regarding use of social media for entertainment, moderate users had 2.832 times higher odds of depression compared to students with low frequency use (p= 0.092). Use of social media to gain information did not show statistically significant differences across any of the groups (p=0.826, p=0.506 respectively). Use of social media to gain information did not show statistically significant differences across any of the groups (p=0.826, p=0.506, respectively). See **(Table 4)** for the complete breakdown of multivariate analysis.

**Table 4:**
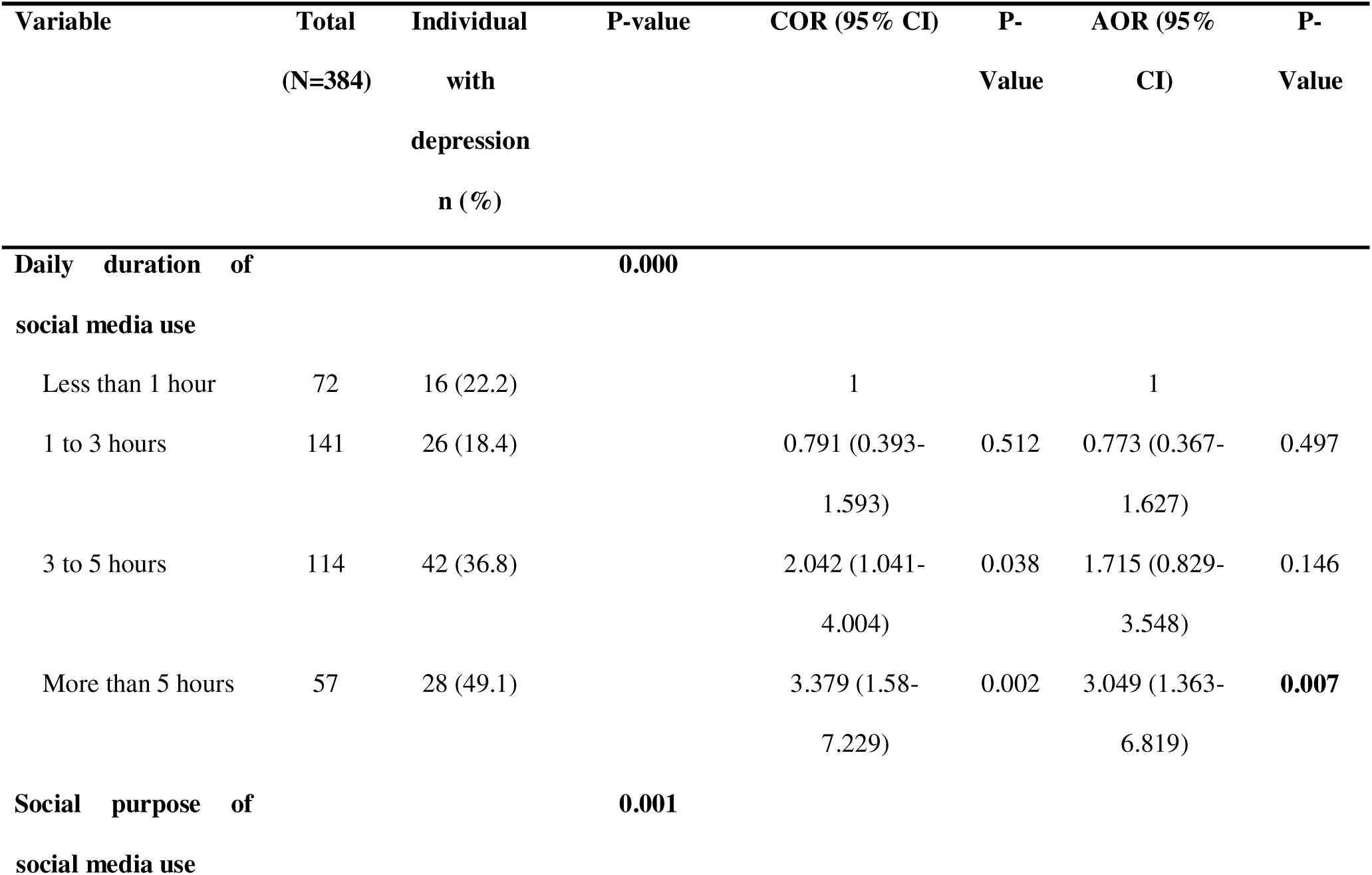
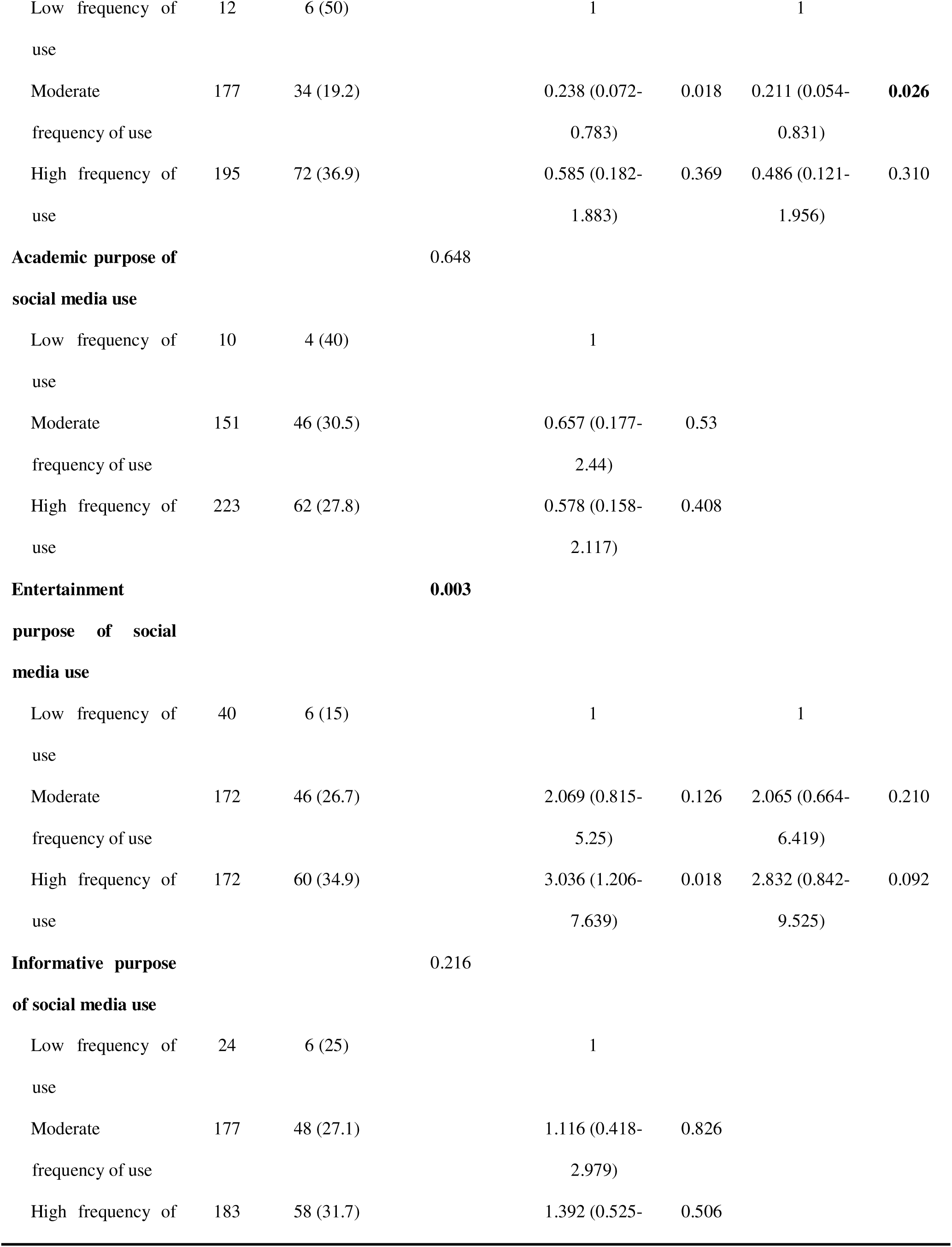
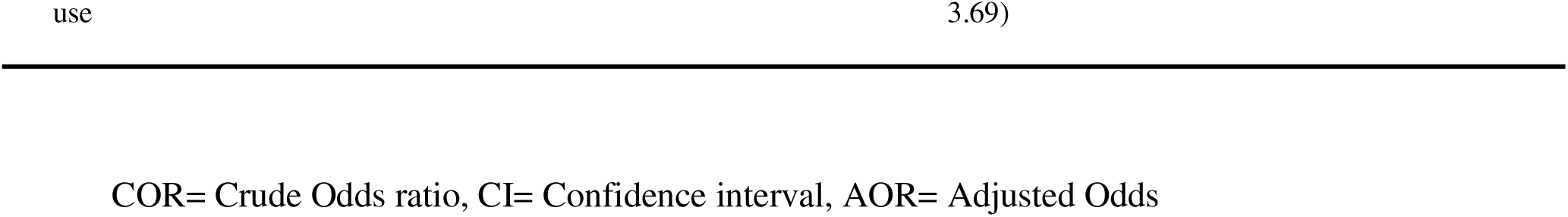
Association between social media usage patterns and depression (N=384).

## Discussion

Social media use and depression are two common phenomena among the university student population. This study aimed to describe the patterns of social media use, and determine the associations between these patterns and the presence of depression among university students in northern Tanzania.

The majority of students on this study reported using social media for 1-3 hours daily. This finding is consistent with a report from Tanzania indicating that social media users spent an average of 1.9 hours on networking platforms and apps (8). Similarly, a study done in Nigeria reported an average daily social media usage of 2-3 hours (33). However, this is contrary to findings of a study from Turkey where only 2% of the sample reported using social media 1-3 hours daily, while the majority reported 4-6 hours of daily use (34). This discrepancy may be attributed to different economic levels and access to technology that may influence social media usage (33,34).

Students in Tanzania may have limited access to internet connections overall because of data costs, and their universities may not offer reliable internet connections (35).

Notably, students who used social media for more than five hours per day on the current study were more likely to experience depression that moderate users. It is possible that students spending more time online had a higher level of depressive symptoms and coped through using social media (36). Alternatively, perhaps the higher-frequency users required more social connection that was missing in their lives, therefore, being more likely to use social media (37).

Our analyses similarly revealed that moderate social media use for socialization was associated with a lower risk of depression, compared to those who used who used it at a lower frequency. This is similar to previous literature from the United States that detected increased levels of depression with low internet use (38). This is a possible indication of the benefits of having an online community and space to socialize. Those who spend less time socializing through social media may be missing a sense of connectedness, therefore, self-reporting higher levels of depression (36).

Our findings also revealed that university students used social media for multiple and concurrent purposes, with the highest reported reason (58.1%) being for academics and learning, while socializing (50.8%), general information (47.7%) and entertainment (44.8%) were also popular reasons. These findings were similar to those from Kenya (11), however, contrasted results from Nigeria where up to 83% of university students primarily utilized social media platforms for socialization (33). The study from Nigeria used a self-developed and previously untested instrument which may explain this difference in findings.

The observed patterns and fairly equal distribution of use of social media for academic learning, socialization, and entertainment suggests that social media universally fills a gap in these needs. Students may prefer online information sources to physical libraries, and overall more passive forms of information seeking (39,40). This is a possible reflection of the natural evolution of access to information over time as internet use increases (39), but also how younger generations view the relevance and significance of different sources of information. In alignment with this, growing literature indicates that educational institutions need to revise how they teach and engage younger learners; mixed-methods may be more effective and preferred compared to traditional in-person teaching (40).

Almost one-third of the university students on this study screened positive for depression; this echoes results from a study conducted in Turkey where a prevalence of 30.3% was detected (34). Previous studies from East Africa revealed a relatively lower prevalence of depression; one from Uganda (16.7%) (17) and another from Tanzania (21.3%) (31). This difference in prevalence across the studies may potentially be driven by differences in methodology and geographical location.

The relatively higher prevalence of depression on this study, and the associations with social media use and depression, indicates that there is a need for raising awareness on these subjects at the individual and institution level (41). National and institutional guidelines exist to inform safe social media use, and the integration of safe use within higher education institutions (42,43). The World Health Organization endorses the Screening, Brief Intervention and Referral to Treatment (SBIRT) framework, originally developed for substance use prevention, as an effective model that can be applied diverse settings in the absence of mental health specialists and other resource limitations (44). Following this model, targeted screening for depression could be paired with brief counselling and awareness for students in need, and refer students for professional support when necessary. Previous studies endorse school social clubs and peer-support as feasible and complementary approaches for settings where mental health services are limited (45,46).

Considering the current findings of one-in-three university students experiencing depression, it is essential for institutions to integrate efforts to screen and support students in need. Studies conducted in the Kilimanjaro region investigating levels of depression in other non-mental health settings, such as HIV care, similarly found a high prevalence of depression with a lack of integrated screening and support for this challenge (47). We therefore, similarly recommend adopting evidence-based guidance from the mhGAP Programme package that offers basic psychosocial interventions that can be conducted by non-mental health specialist individuals (48). Coupled with the SBIRT model, support staff could be empowered to identify and support students experiencing these common challenges (44).

Although the TCRA offers regulations on appropriate internet and social media usage, the terms of safe use may be poorly understood at the community level. Further, the landscape of social media use is evolving quickly; youth may face difficulties in maintain constant vigilance and rationalization while accessing overwhelming amounts of new information on social media. Both students and staff within universities should receive regular refresher training on how to safely engage with online information sources and the risks of social media use (43,49).

This study has several limitations. The cross-sectional design of the current study limits the ability to establish causality. While associations can be identified, it remains unclear whether social media use contributes to depression or if students experiencing depression are more likely to engage in excessive social media use.

Additionally, we acknowledge that reliance on self-reported data may introduce responder and social desirability biases, such as underreporting or overestimating social media use and depression. These biases are common in survey-based mental health research and have been documented in previous studies (50,51). While sharing the survey with participants, we attempted to minimize the risk of bias by asking participants to respond honestly and by reassuring anonymity of their data.

Some of the tools utilized on this study have not been validated for use in the current context or tested for cultural appropriateness, therefore, we encourage readers to be mindful of this while interpreting findings. Our team’s efforts to pre-test the tools for comprehension hopefully mitigates some of the potential bias caused by this.

## Conclusion and recommendations

Prevalence of depression was high in this setting. Daily duration of social media use, of five hours or more, was significantly associated with depression. Moderate socialization through social media was negatively correlated with depression. These findings indicate the utmost importance of raising awareness on the potential benefits and harms of social media use, at both the individual and institution level. We further recommend integration of screening and support programs for students in need (45,46). Ultimately, both students and educational institutions both have roles to play in maintaining good mental health in relation to the use of social media (43).

## Data Availability

Deidentified datasets used and/or analysed during the current study are available through URL upon request from Beatrice Temba at. BT is a non-author and ethics committee member at KCMC University.

## Acknowledgements

Not applicable.

